# Healthcare practices for individuals with cerebral small vessel disease: An International Survey

**DOI:** 10.1101/2025.06.03.25328926

**Authors:** Carmen Arteaga-Reyes, Joanna M. Wardlaw, Fergus N. Doubal

**Author notes:** Correspondence to: Fergus N. Doubal, University of Edinburgh, Chancellor’s Building, 49 Little France Crescent, Edinburgh, EH16 4SB, UK.

## Abstract

**Background:** Cerebral small vessel disease (cSVD) is a major cause of stroke and dementia but patients often report a lack of specialist care. We aimed to identify the availability and type of care, current management practices and areas of unmet need for patients with cSVD.

**Methods:** We performed a literature review to identify clinical practices towards cSVD and an online international survey of health-care centres (one clinician per hospital/clinic) treating individuals with cSVD between September/2022–May/2023. All answers were anonymised (with an option to sign). We compared the clinical workup between cSVD-dedicated and non-cSVD-dedicated health-care services, regions and country-income-class groups using descriptive statistics.

**Results:** Our review found five clinical cSVD-services. We distributed 264 survey requests and received 137 responses (response rate=52%); 130 responses representing 45 different countries contained analysable data; 63% responses were from High-Income countries (HIC). Fourteen centres/130 responses (11%) had cSVD-services, nine in HIC, seeing a median of 150 (IQR 64.5,290) individuals with cSVD/year. 79% (91/115) physicians reported cognitive decline as the primary concern for patients. Follow-up was more likely in cSVD-services (cSVD-centres vs not: 85% 11/13 vs 45% 49/109; ꭕ^2^(df)=1, p=0.007) for main outcomes stroke and dementia. 77% (84/109) centres without a cSVD-service believed there is an unmet clinical need for cSVD. 21% did not assess cognition, and 21% (25/117) applied diagnostic cognitive tools. For covert-cSVD, antiplatelet use was more likely in Latin-America & the Caribbean (Fisher-exact, p<0.001) and statins in Europe & Central Asia (Fisher-exact, p=0.01). 61 services evaluated long-term outcomes, of which 8 reported patient-reported-outcome measures.

**Conclusions:** This survey has identified very few cSVD specialist services, a large, physician-acknowledged unmet clinical need, and serious mismatch between clinical management and patient-reported priorities for people with cSVD. Our results indicate a need for improved person-centred care for cSVD. Standardisation of practices and services could significantly improve health-care for people with cSVD.

## INTRODUCTION

Cerebral small vessel disease (cSVD) is a leading cause of stroke, dementia and multi-systemic symptoms.^1^ Guidelines relevant to cSVD management exist, ^2–5^ however the availability of services for individuals with cSVD is unclear and persons affected by cSVD report unmet clinical needs.^6^ Additionally, previous studies suggested that a coordinated approach to care for individuals with cSVD may help to tackle the existing gaps in their care overlooked by a traditional stroke/dementia-care approach.^6–8^

Patients with cSVD present to different specialities and can fall between services, experiencing fragmentation of care leading to a failure to convey the diagnosis, information about prognosis and a lack of management which may lead to worse clinical outcomes.

We hypothesised that there are few health-care services dedicated to individuals with cSVD, a lack of standardised (written or unwritten) clinical practices amongst clinicians managing people with cSVD, and that there are critical gaps in current care for individuals with cSVD.

We performed a literature review to identify health-care practice patterns for individuals with cSVD. We also designed and performed a survey which aimed to: 1) Identify cSVD-dedicated clinical services; 2) Investigate current clinical practices, attitudes, perceptions, and experiences of clinicians managing individuals with cSVD, including person-centred care practices; 3) Identify potential gaps in clinical management; and 4) Identify whether differences in practices are related to the type of clinical service, geographical regions, or economic classification of the participating countries.

## METHODS

### Literature review

We searched Ovid EMBASE and PubMed for published literature in Spanish, English or French to identify: 1) cSVD-services and their clinical practices, and 2) previous surveys investigating cSVD practice patterns. We searched from inception to April 9^th^, 2025; supplemented with searches of reference lists of identified papers. We used the terms “cerebral small vessel diseas*.mp.” and “(attitude* or practice patterns or healthcare practice or experience practice or practice management or gaps or delivery of healthcare).mp.” or (survey or questionnaire).mp.”. We screened 2,795 papers and collected data from relevant papers (Figure S1).^9^

### Design and Data collection

We designed our quantitative online survey for practising clinicians managing individuals with cSVD based on current guidance on survey design^10,11^ and on care for individuals with cSVD.^3–5,12–18^ Data were collected using SurveyMonkey Inc. The survey was developed by consensus between a vascular neurologist (CAR), stroke and geriatrician clinician (FND) and neuroradiologist (JMW) with expertise in cSVD. We pilot-tested our survey on our local Lothian NHS cerebrovascular clinicians’ team (15 clinicians, including staff with expertise in survey design), who were representative of our target population. Ambiguous items were amended, correct question logic was revised, neutral wording was carefully used to prevent interviewer bias, and the current survey version was finalised. We report our survey based on the Checklist for Reporting Of Survey Studies (CROSS) recommendations.^19^

Clinicians responded to a subset of 59 questions with estimated completion time of 10 minutes. The survey was available in English language only (Supplementary material–Survey Full-text). To minimise incomplete responses, we used skip and logic functions based on five main questions: 1) Background specialty (neuroradiology vs other clinical specialties); 2) Type of current clinical service of practice (cSVD-dedicated service vs non-cSVD-dedicated service); 3) Clinician’s perception of an unmet clinical need for individuals with cSVD in their region (‘Yes/No/Don’t know’); 4) Clinician’s perception of a cSVD-dedicated service usefulness (‘Yes/No’); and 5) Routine follow-up of individuals with cSVD offered by clinician (‘Yes/No’).

The survey collected demographic characteristics from all clinicians. Then, based on the question tree, clinicians were asked questions tailored to their type of service. Most of the questions allowed multiple answers, and multiple option questions included ‘none/not applicable’, ‘other (specify)’ with free text sections.

We used “clinically overt cSVD” for stroke, cognitive, mobility and balance syndromes, with a separate category for suspicion of genetic causes of cSVD (’possible’). We used the category “moderate-severe covert-cSVD found on scanning for other purposes” to refer to individuals presenting without overt cSVD-syndromes in whom neuroimaging hallmarks of cSVD were present. We selected the umbrella term *‘covert*’ versus *‘asymptomatic/silent/incidental’* to reflect the current common term used in the cSVD literature to capture individuals *‘truly asymptomatic’* and with incidental cSVD-radiological biomarkers. We employed the option “severe cSVD (however it is defined)” to capture clinicians’ attitudes based on cSVD-severity on imaging as opposed to clinical presentation.

We investigated availability of person-centred care and health-care support. We asked clinicians about nine main potential areas of concern expressed to them by individuals with cSVD based on: a) Major clinical syndromes associated with cSVD;^1^ b) Previous reports of common concerns by people living with cSVD;^6^ and c) The authors’ clinical and research experiences. We asked if standard patient information (any format) were provided by their clinical service. Finally, we asked about patient or relative referrals to other relevant support (e.g., voluntary health organisations).

### Sample characteristics and Survey administration

We distributed our online survey (30^th^/September/2022–31^st^/May/2023) for practising vascular brain clinicians identified from the World Stroke Organisation (WSO), European Stroke Organisation (ESO), The International Society of Vascular Behavioural and Cognitive Disorders (Vas-Cog), and the Ibero-American Stroke Organisation (IASO). We complemented our list of invitees with clinicians from relevant papers and used a ‘hospital-limited’ snowball sampling method.^20^ The survey targeted one health-care clinician per centre managing individuals with cSVD. All answers were anonymised, with an option to sign. Responses were treated confidentially and data were only accessible to authorised researchers. Online survey was closed on 4^th^/June/2023.

Invitations to complete the survey were delivered via general/impersonalised email, including instructions and researchers’ contact details. To maximise participation, reminders were sent at one and two-months. To prevent multiple participation, we limited to a single open attempt of the survey, with option to contact researchers if difficulties while completing the survey. Duplicated responses from clinicians experiencing an unforeseen technological difficulty were addressed and responses were excluded from analysis (n=5).

### Statistical analysis

We downloaded final survey data on 9^th^/June/2023. We used descriptive statistics (Chi-squared, Fisher’s-exact, Mann-Whitney-U) reporting individual P-values as appropriate and report frequencies for categorical variables and median (interquartile range, IQR). We assessed relationships between types of services (cSVD and non-cSVD), between geographical regions, and between country income class, using the World Bank region and income classification.^21^

We defined ‘invitees’ as all who received an email invitation, ‘respondents’ as all who had opened the survey, ‘complete responses’ as answers provided to all questions, and ‘partial responses’ when analysable data was provided but incomplete. Responses without analysable data (n=2, unit of non-response) were considered as ‘missing data’ and approached as listwise deletion. Partial responses with analysable data were included. All analyses were performed with IBM SPSS Statistics v27 and figures were created using R 4.2.764 (packages ggplot2 and maps).^22–25^

### Ethics

This survey did not require ethical review and approval.^26^ Agreement to participate in the survey was taken to indicate consent.

### Data Sharing

Data are available from the corresponding author upon request.

## RESULTS

### Literature review

#### cSVD-services and their clinical practices

We found five cSVD clinical services, (key findings in Table S1).^27–34^ Multidisciplinary and structured care were the standard. Three of five services provided care to individuals with sporadic and genetic cSVD.^30–32,34^ The other two were dedicated to individuals with cerebral amyloid angiopathy,^27,33^ and CADASIL.^28,29^

#### Surveys investigating healthcare practices for individuals with cSVD

We found six surveys, using different survey tools (n=5/6)^28,35–38^ and semi-structured interviews(n=1/6),^8^ (Table S2). Previous surveys assessed: 1) uncertainties and management of covert brain infarcts;^8,35^ 2) suitability of remote follow-up for people with CADASIL;^28^ 3) reasons for diagnostic choices made by individuals at risk of CADASIL;^36^ 4) emergency clinicians’ practices towards transient ischaemic attack (TIA) or minor stroke;^37^ and 5) brain magnetic resonance imaging (MRI) reporting practices in individuals with cognitive disorders, including people with cerebrovascular disease.^38^ We did not find surveys investigating clinicians’ practices in the management of the cSVD-spectrum, whether at cSVD-or non-cSVD-services.

### Survey results

#### 1. Respondents’ demographics

##### 1.1 Response rate

We received 137 responses from 264 invitations (response rate 52%). Each response represented one clinician managing individuals with cSVD per clinical health-care service, from 45 different countries in five continents, (Figure 1). We excluded five duplicated responses (identified through direct contact with clinicians); we then used the most complete response. We also excluded responses with no analysable data; total excluded n=7/137, leaving 130/137 responses with analysable data, (Figure 2). Denominators changed across the results due to the skip and logic functions, the multiple answers option in the questions (see methods); and the partial responses with analysable data.

**Figure 1.**
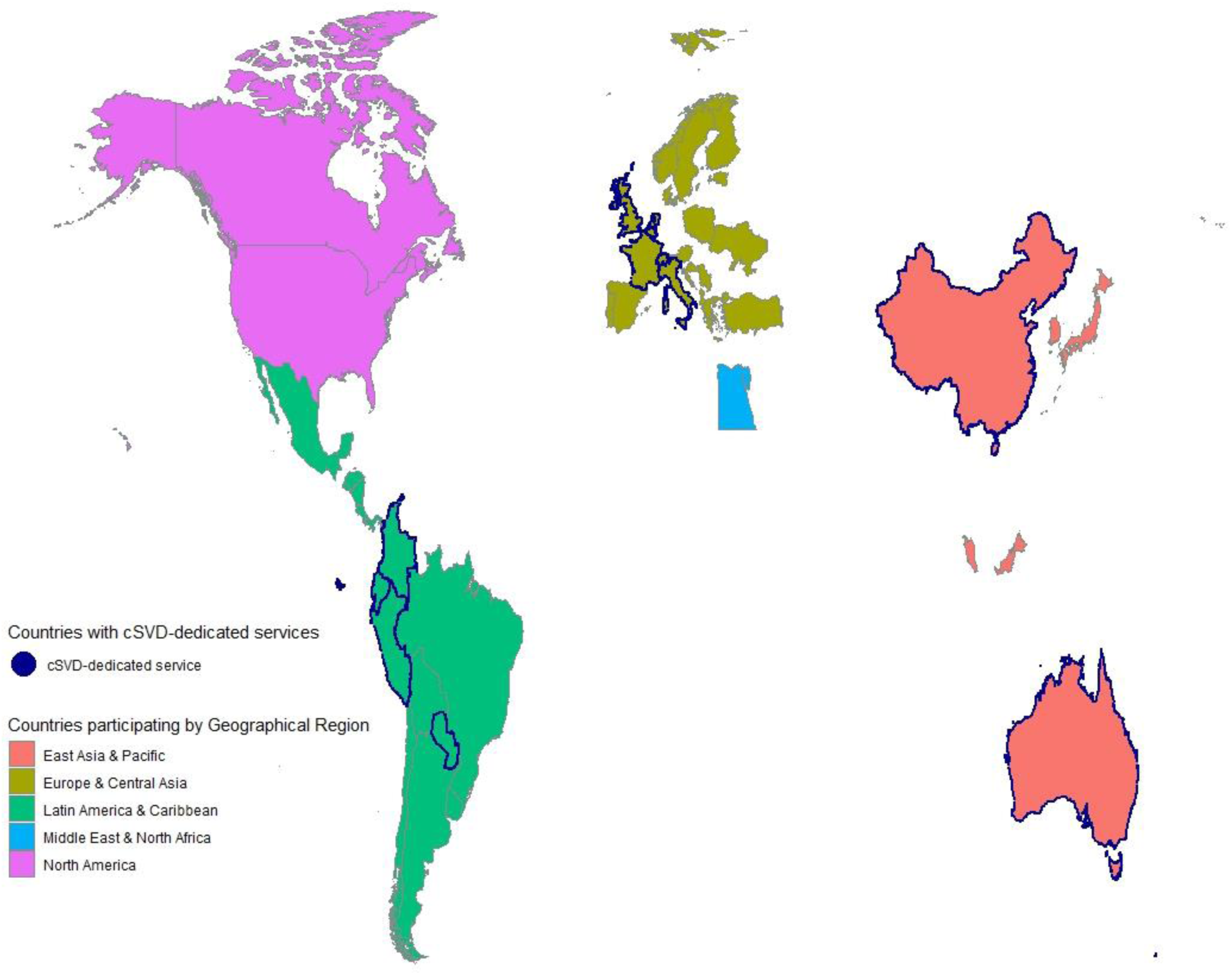
Respondents’ country of practice and cSVD-services Responses involved 45 countries within five geographical regions. cSVD-dedicated-services identified (n=14) localised in Australia, Belgium, China, Colombia, Ecuador, France, Italy (2/14), Netherlands, Paraguay, Peru, Switzerland, United Kingdom (2/14). cSVD-Cerebral small vessel disease.

**Figure 2.**
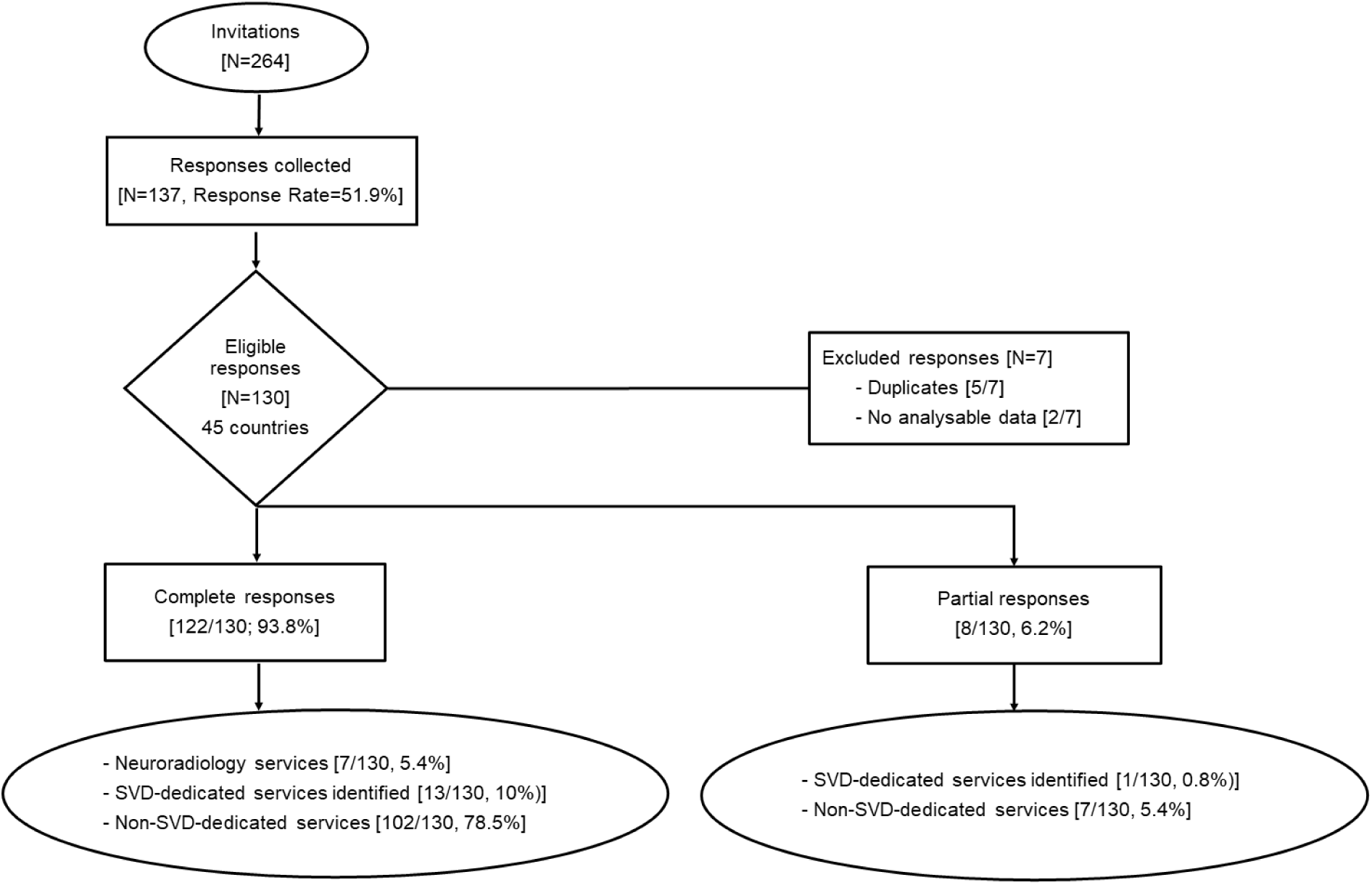
Flow chart responses *Duplicates–Duplicate responses from same clinician. ^†^No analysable data–Only country, specialty and years of practice available.

##### 1.2 Response sources

Responses were mainly from Europe & Central Asia (ECA; 71/130, 55%) and Latin America & Caribbean regions (LATAC; 42/130, 32%). Despite active attempts of surveying the remaining regions, we only collected 1/130 response from the Middle East & North Africa (MENA), (Figure 3). Responses came, in order of frequency, from High- (HIC), Upper-Middle (UMIC) and Lower-Middle (LMIC) income countries (63%, 33% and 4%, respectively), with null representation from Low-income countries (LIC).

**Figure 3.**
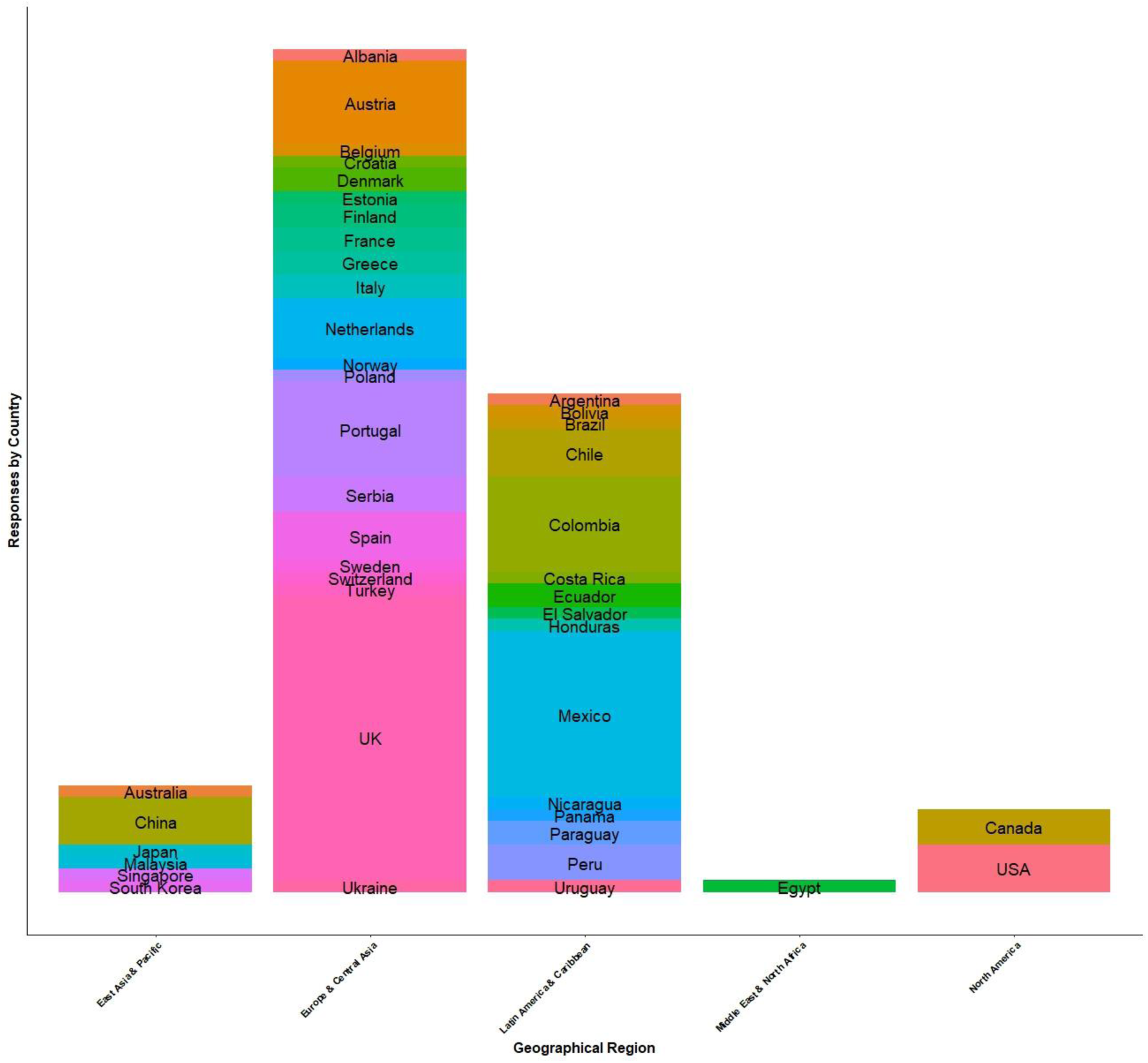
List of participating countries per region East-Asia & Pacific (7%); Europe & Central-Asia (55%); Latin-America & Caribbean (32%); Middle-East & North-Africa (1%); and North-America (5%)

##### 1.3 Clinicians’ background

95% (123/130) of respondents were vascular brain specialists, and 5% (7/130) had a neuroradiology background.

#### 2. Clinical service demographics

We identified 14/123 (11%) cSVD-services. 8/14 were located in ECA region, and 9/14 in HIC. 77% (84/109) of clinicians without a cSVD-dedicated service perceived a local unmet clinical need for individuals with cSVD, and 87% (73/84) considered a cSVD-dedicated service would be useful.

##### 2.1 Neuroradiology services (n=7/130)

Three neuroradiologists (3/7) reported having a standard imaging and reporting protocol for detecting cSVD findings on brain scans. 4/7 neuroradiologists routinely used scores to assess cSVD load (‘burden’) on brain scans (most commonly Fazekas, 3/7). Three neuroradiologists had standard protocols to directly refer individuals with ‘moderate-severe symptomatic or covert cSVD’.

##### 2.2 cSVD-services

cSVD-services (14/130) saw a median of 150 (IQR 64.5, 290) individuals with cSVD/year. 81% reported referrals from three specialty services (Neurology/Stroke-TIA/Memory clinics), 57% also from general practitioners.

All 14 cSVD-services were led by a vascular brain specialist; half of them (7/14) had at least one mental health professional (Psychiatrist/Psychologist/Neuropsychologist); and six had a physiotherapist and/or occupational therapist; providing multi-disciplinary care for individuals. Only one cSVD-service reported nursing, social worker, and dietician in their team

#### 3. Comparisons between cSVD-dedicated and non-cSVD-dedicated services

##### 3.1 Who is/should be seen in cSVD-dedicated clinics?

All 13/13 (data from one centre incomplete) cSVD-services saw individuals presenting with cognitive impairment. Additionally, 10/13 (77%) saw patients with stroke, genetic causes or moderate to severe covert-cSVD. 8/13 (61%) also saw patients with mobility issues and one clinic focussed on cerebral amyloid angiopathy (CAA)/young cSVD.

In centres without cSVD-services, 82% (60/73) of clinicians that considered a cSVD-service would be useful believed all groups of individuals with cSVD, i.e., stroke/genetic/mobility/movement-related disorders/moderate–severe covert cSVD presentations, should be seen in a cSVD-service. Cognitive presentations were reported as a clinical priority 93% (68/73), (Table 1). However, 33% (36/109) believed none of these groups of individuals with cSVD should be seen in a specialised cSVD-service as their needs were believed to be met in existing services.

**Table 1.**
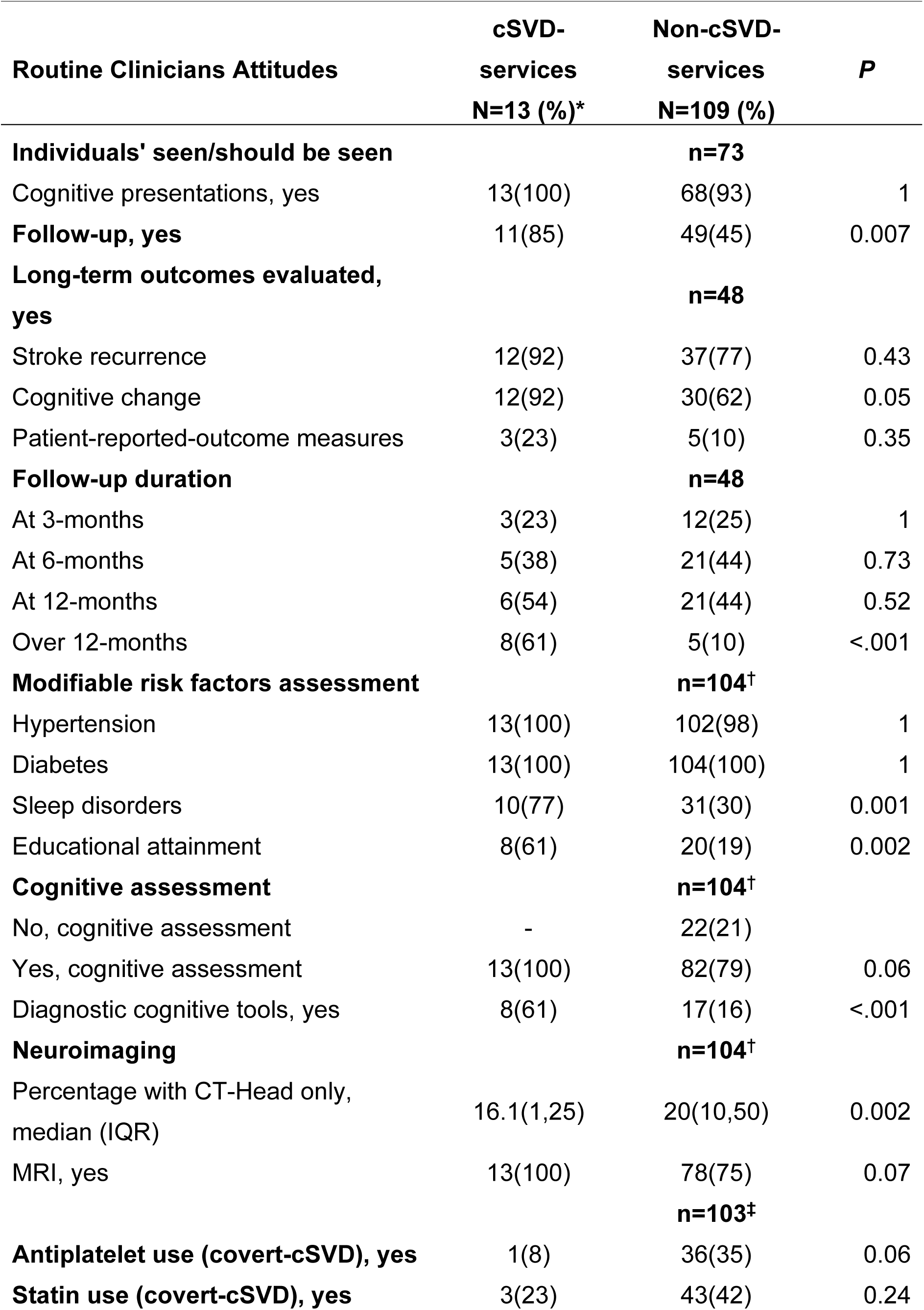
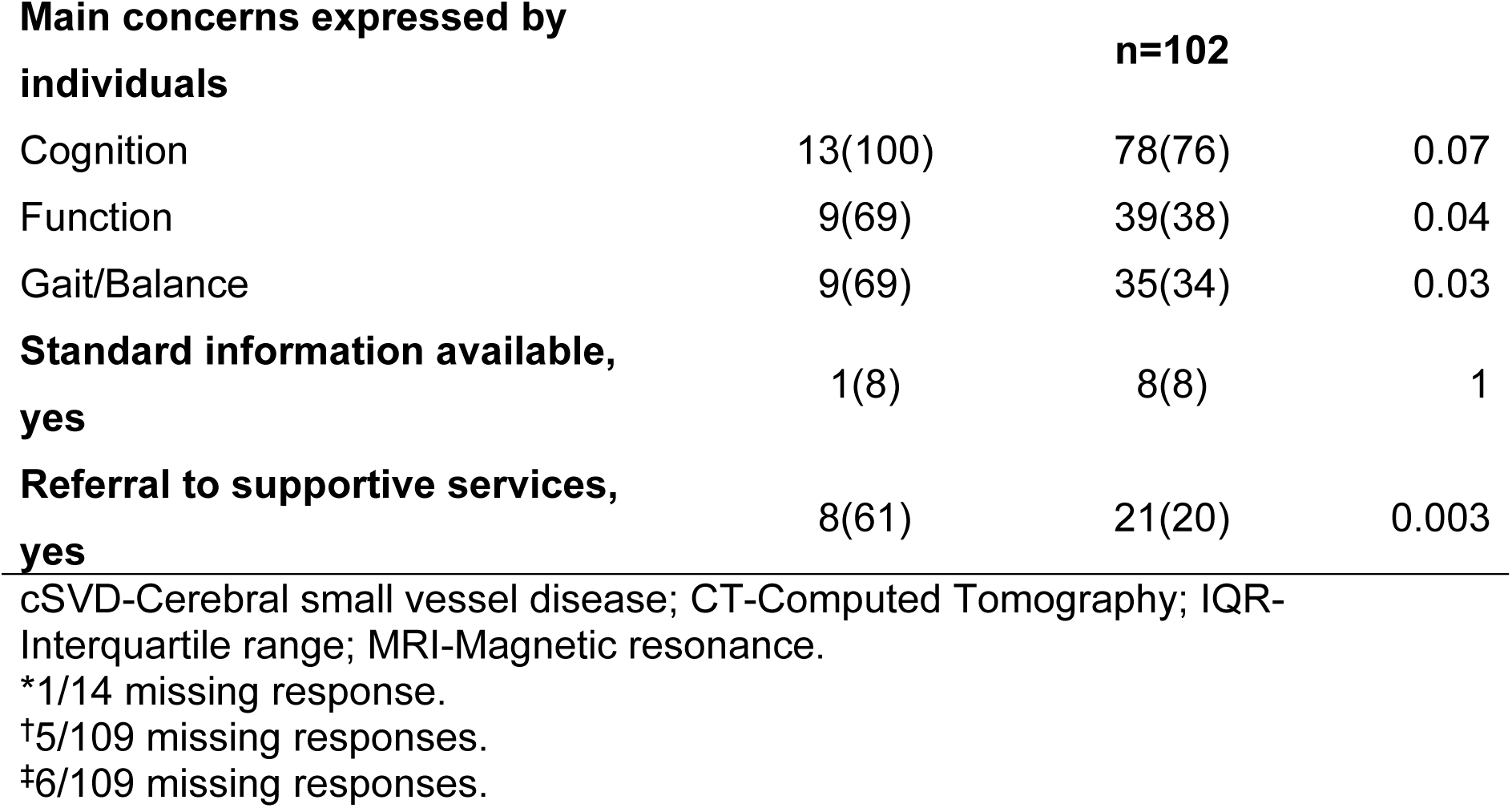
Service differences between cSVD-and non-cSVD-centres.

##### 3.2 cSVD patient follow-up?

cSVD-services routinely followed-up a higher proportion of patients with cSVD than non-cSVD-services (85% 11/13 centres vs 45% 49/109 respectively; p=0.007). We found that both types of service tended to follow-up clinically overt cSVD (stroke/cognitive/mobility) presentations (cSVD-services 71% vs non-cSVD-services 43%; p<0.001), while covert-cSVD is the least likely to be followed-up (cSVD-services 54% vs non-cSVD-services 16%; p=0.02). Additionally, individuals with cSVD and uncontrolled risk factors were followed-up in 61% of cSVD-services and 30% of non-cSVD-services, p=0.02.

##### 3.3 Long-term outcomes (What and When)

Amongst respondents who offered routine follow-up, stroke recurrence and change in cognition were more commonly assessed in cSVD-services 92% (12/13) vs non-cSVD-services 62% (30/48); p=0.05. Patient-reported outcome measures (PROMs) were the least common reported outcomes measured (23% 3/13 cSVD-services vs 10% 5/48 non-cSVD-services). In cSVD-services single follow-up was the most common practice, while no follow-up assessment at all was the main practice in non-cSVD-services.

##### 3.4 Routine clinical assessment in individuals newly diagnosed with cSVD

We explored clinicians’ ‘compliance’ to basic general aspects of primary and secondary care that should be assessed in individuals with cSVD as per current clinical guidelines.^4,5,13,16,17,39,40^

###### Risk factors

Diabetes (100%), hypertension (98%) and smoking (97%) were the main risk factors assessed across centres, followed by lipids (96%), alcohol intake (77%), anthropometric measures (53%), and physical activity (51%). Diet (i.e., nutrition) was assessed in 49%, and other modifiable risk factors (modifiable-RF) were seldom reported (i.e., hearing/vision impairment 0.8%). Less *‘traditional’* modifiable-RF, i.e., sleep disorders, were assessed more frequently in cSVD-services (77% vs 30% non-cSVD, p=0.001). 61% of cSVD-services and 19% of non-cSVD-services assessed educational attainment (p=0.002), but only 6% of all services reported childhood deprivation assessment.

Additionally, there appeared to be a trend to higher use of antiplatelets and statins in individuals with covert-cSVD in non-cSVD-services. 7% (1/13) cSVD-dedicated services routinely prescribe antiplatelets and 35% (36/103) in non-cSVD-services (p=0.03). Similarly for statins, 3/13 (23%) and 43/103 (42%) respectively, (Table S3).

###### Examination

Complete neurological exam was performed in 92% (12/13) cSVD-services and 89% (93/104) non-SVD-services. However, formal assessments of specific examination components were inconsistently performed in both settings: functional (cSVD-services 69% vs non-cSVD 55%); mood/behaviour (38% vs 21%); dexterity/mobility/motor (23% vs 8%); health-related quality of life (0% vs 7%); and information retrieval from relatives was uncommonly performed (29% vs 5%; p=0.01).

###### Cognitive tests

In 21% (22/104) of non-cSVD-services cognition was not routinely assessed in individuals newly diagnosed with cSVD (vs 100% 13/13 in cSVD-services).

Of those reporting assessing cognition there were 17 different cognitive functioning tools, (Table S4). Interestingly, only 31% (4/13) of cSVD-services performed neuropsychological assessment (vs 6% 6/104 of non-cSVD-services, p=0.01). 80% (94/117) of clinicians across centres use Montreal Cognitive Assessment (MoCA) in clinical practice, followed by Mini-Mental State Examination (MMSE) in 53% (62/117).

Only one clinician (1%) reported using the National Adult Reading Test (NART) at a non-cSVD-service; no additional tools for premorbid cognition were reported.

###### Investigations

Overall, most clinicians routinely performed hypertension (98%, 112/117) and diabetes screening (95%, 111/117, any test), and 78% (91/117) included electrocardiogram (ECG) in individuals with newly diagnosed cSVD. Other routine investigations were kidney function 49% (57/117) and retinal examination 9% (11/117). Only 3% (4/117) of clinicians in non-cSVD-services did not perform routine investigations.

###### Neuroimaging

We explored the use of neuroradiology for individuals newly diagnosed with cSVD. 100% (13/13) of cSVD-services and 75% (78/104) of non-cSVD-services ensured that individuals with cSVD have a recent MRI, (p=0.03). Overall (n=117), FLAIR, DWI, haemosiderin-sensitive (T2*–W and/or SWI), T2-W, and T1-W sequences were justified in 89%, 85%, 84%, 80%, and 76%, respectively, (Table S5). A greater proportion of individuals seen in non-cSVD-services had CT-Head only (median=20; IQR [10, 20]) vs cSVD-services (median=16.1; IQR [1, 25]; z=-3.04, p=0.002). See other neuroimaging tests reported in Table S6.

##### 3.5 Routine person-centred care and health-care support Personalised care – Person’s concern expressed to clinicians

Cognition was the main patient-reported concern followed by (lack of) treatment and prognosis (Table 2).

**Table 2.** Concerns expressed by individuals with cSVD to clinicians.

| **Concern related to cSVD (in order of frequency)** | **All services**<br>**N=115 (%)** |
| --- | --- |
| Cognition or dementia risk | 91 (79) |
| cSVD-Treatment | 65 (56) |
| Risk of Stroke/TIA occurrence OR recurrence | 64 (56) |
| cSVD-Diagnosis | 61 (53) |
| Risk factors and their management | 61 (53) |
| Functional impairment | 48 (42) |
| Gait/Balance disorders | 44 (38) |
| Mood/Affect disorders | 27 (23) |
| Urinary disorders (urgency/incontinence) | 23 (20) |
| None/Not applicable | 2 (2) |
| Other | 4 (3) |
| Prognosis | 2 (2) |
| Aetiology (especially in none/low cardiovascular risk factor individuals) | 1* |
| Heredity | 1 |
| Effective interventions to halt cSVD progression | 1* |

cSVD-Cerebral small vessel disease; TIA-Transient ischaemic attack.
\*Two items, one clinician's response

Concerns related to functional impairment (cSVD-service 69% vs non-cSVD-service 38%, p=0.04) and gait/balance disorders (69% vs 34%, p=0.03) were reported more frequently by clinicians in cSVD-services versus non-cSVD.

###### Empowering patients – Standard information available for individuals with cSVD and their relatives

Standardised information (any format) was lacking, with only one cSVD-service and eight non-cSVD-services reported having standardised information for people with cSVD (9/116, 8%), in seven countries and two regions, ECA (7/9) and LATAC (2/9).

###### Supportive care – Referral of patients

61% (8/13) of clinicians in cSVD-services routinely refer individuals with cSVD to supportive services, including but not limited to voluntary health organisations, community services and other clinical services for cSVD-related symptoms. While in non-cSVD-services referrals were reported for 20% (21/103, p=0.003).

## DISCUSSION

This systematic review and large international survey with a good response rate and reasonable geographic spread (45 countries, five continents) has demonstrated a lack of specialist services for patients with cSVD. Furthermore, there is wide variation in assessments and significant heterogeneity in management of patients with covert-cSVD. Despite this, most (82%) clinicians in non-specialist cSVD services acknowledge an unmet clinical need for individuals with cSVD, which concurs with patients who report cognitive concerns.^6^

Overall, our literature review highlighted that significant advances in the understanding, identification, and management of individuals with cSVD have not yet translated to clinical practice. Previous surveys performed at non-SVD-dedicated services showed lack of consistency in multidisciplinary communication, underreporting, prognosis uncertainty, and scarcity of institutional written protocols for care of people with cSVD.^8,35,37,38^ However, there were relevant publications describing evidence-based clinical services for cSVD^27–32,34^ established on the initiative of a few motivated individuals, that may be beneficial to individuals with cSVD in individual local circumstances.

Our survey found that most of the cSVD-centres were currently located in the ECA region (8/14) and in HIC countries (9/14). Yet, the perception of unmet clinical needs for people with cSVD was global. The median of 150 (IQR 64.5, 290) individuals with cSVD/year seen at cSVD-services is insufficient considering the global burden of cSVD causing stroke and dementia.

Our results showed that even in specialised services (e.g., cerebrovascular services), management varies, from assessment of modifiable-RF (where we can prevent/act) to more complex assessments such as diagnostic cognitive assessment or neuroimaging (what it is helpful to know). Strikingly, our results also revealed that assessments related to functional, physical, neurobehavioural, and health-related quality of life were scarcely performed in both settings, cSVD and non-cSVD-services. In addition, there was little assessment of early life factors, adverse social determinants of brain health (e.g., “childhood deprivation”, or education attainment), and other risk factors for cognitive impairment or dementia (e.g., hearing/vision impairment).^41,42^

Previously published data from a SVD–centre, found that post-stroke and incidental (“covert”) cSVD-related neurobehavioural symptoms were the main reasons for referral to their service (89%, N=525/587), diagnosing vascular mild cognitive impartment/dementia 17% of their patients.^34^ However, our results showed that while cSVD cognitive symptoms and outcomes were widely recognised as a major concern of patients and health-care priority across sites; 1 in every 5 individuals with cSVD attending non-cSVD-services would not receive routine cognitive assessment. Furthermore, cognitive assessment in the acute post-stroke phase has been described as key to managing early rehabilitation.^39^ We found 17 different cognitive-functioning tools clinicians use, with a higher variability among those in non-cSVD-services. This exhibits the heterogenous use of screening and diagnostic cognitive functioning tools in clinical settings.

Our results identified that only 1 in 2 non-cSVD-services (most available type of clinical service) followed-up individuals with overt-cSVD, and almost 2 in 10 centres would follow individuals with covert-cSVD. An implication of these findings is the possibility that early identification of individuals “converting” from truly asymptomatic to symptomatic (apart from stroke symptoms) is unlikely to be captured at specialty services. Underscoring the urgent call for education, sensitisation and standardisation of care for people with cSVD, starting from primary care.

Overall, pharmacological secondary management of overt ischaemic cSVD-stroke has been traditionally based on two pillars: modifiable-RF control (including use of lipid-lowering drugs), and antithrombotics (mostly antiplatelets). Worryingly, antiplatelet drugs may be more likely to be used in covert cSVD in non-specialist than in specialist cSVD services (Table S3) despite potential evidence of harm in one guideline,^2^ underpinning a need for more specialist service provision for cSVD.

We explored practices towards detecting modifiable-RF (including “Life’s essential 8”).^43^ We found a higher proportion of assessment of BP, blood glucose, smoking, and cholesterol, whereas body mass index, physical activity and diet-alcohol were assessed by half of the clinicians. Regarding neuroimaging assessment, as we anticipated, a greater proportion of individuals with cSVD seen at non-cSVD-services had CT-Head only. While CT-Head is highly useful, it does not assess the complete spectrum of cSVD hallmarks; which have a prognostic value to identify individuals at higher risk for stroke, cognitive decline and mortality.^44,45^ Our results did not explore differences in assessments between the heterogeneous clinical presentations of cSVD; we explored routine assessments in individuals with new diagnoses of cSVD.

It is concerning that services do not routinely follow-up patients with non-controlled modifiable risk factors although this may be devolved to other specialities, e.g., general practitioners. Previous studies have suggested that this conventional practice at stroke clinics may prevent long-term improvement in modifiable-RF control.^46^ Regarding antiplatelet prescriptions for individuals with covert-cSVD when no other indication existed, we found regional and country-income-class differences attributed to higher use of antiplatelets in LATAC and UMIC countries. In contrast, the use of statins in people with covert-cSVD without other medical indications was higher in ECA region. Currently, there is no strong evidence to support the use of these two drugs for this population.^2,47^ One possible explanation for this observation might be differences between MIC and HIC in access to pharmacological treatments and regional regulations. Our findings also align with variability in clinical practices, previously described.^8,35^

Cognitive symptoms/Dementia risk were reported by clinicians as the main concern expressed by individuals with cSVD (79%) consistent with findings of Hwai et al.^6^ We also found amongst centres offering routine follow-up that only 8/61 assessed PROMS. This underlies deficient person-centred care protocols in health-care services. Another interesting finding was that formal information retrieval from relatives was scarcely reported. cSVD has subtle symptoms that can be overlooked by individuals, including early cognitive/behavioural symptoms. Furthermore, informant-reported cognitive complaints have been related to functional impairment and WMH-severity in individuals with covert-cSVD.^48^

Our survey has several limitations; it was designed in English, online access, and invitation-based only, limiting its administration and accessibility, and likely has selection bias. The distribution list was mainly identified from neurovascular disorders organisations; practices in non-neurovascular services, e.g., general neurology or general medicine, may be even more lacking. The small number (8/130) of incomplete responses limited sensitivity analysis. Our sample lacks representativeness of specific regions and income-country-class. However, we succeeded in achieving responses from 45 countries; and we highlighted the need for well-established communication strategies (including low-income countries). Although our multiple-choice questions provided specific test options, we included free text fields to capture the heterogeneity of assessments/tools. Our analyses were limited by the small number of cSVD-services identified/available. We did not investigate referral pathways and staff types working at non-cSVD-services. This approach was adopted since we presumed higher variability in the referral services that would have been otherwise challenging to identify by respondents in higher volume services.

While there are surveys addressing healthcare practices related to cSVD (e.g., management of CBI)^8,28,35–38^ they do not provide a comprehensive overview of clinical practices and attitudes towards individuals with cSVD-spectrum. Our results showed patterns of practices, gaps and challenges faced by clinicians providing care for individuals with cSVD-spectrum. Although the challenges and lack of specific pharmacological treatments for people with cSVD may seem discouraging, we believe that much of this burden could be reduced by standardising clinical processes, rigorous application of known guideline-based risk factor management, inter/intra-disciplinary collaboration, efficient awareness/training, and communication strategies that hit general public and health-care providers.

## CONCLUSIONS

This literature review and large, international survey with a good response rate and reasonable geographic spread (45 countries) has demonstrated a lack of specialist services for individuals with cerebral small vessel disease. Furthermore, there is a variety of assessments and significant heterogeneity in management of patients with covert-cSVD. Most clinician respondents recognised an unmet clinical need for people with cSVD, echoed by the patients themselves. Standardisation of clinical health-care protocols and clinicians’ skills for identification, assessment, reporting and management may improve long-term outcomes for individuals with cSVD.

## Data Availability

Data are available from the corresponding author upon request.

## Non-standard Abbreviations and Acronyms

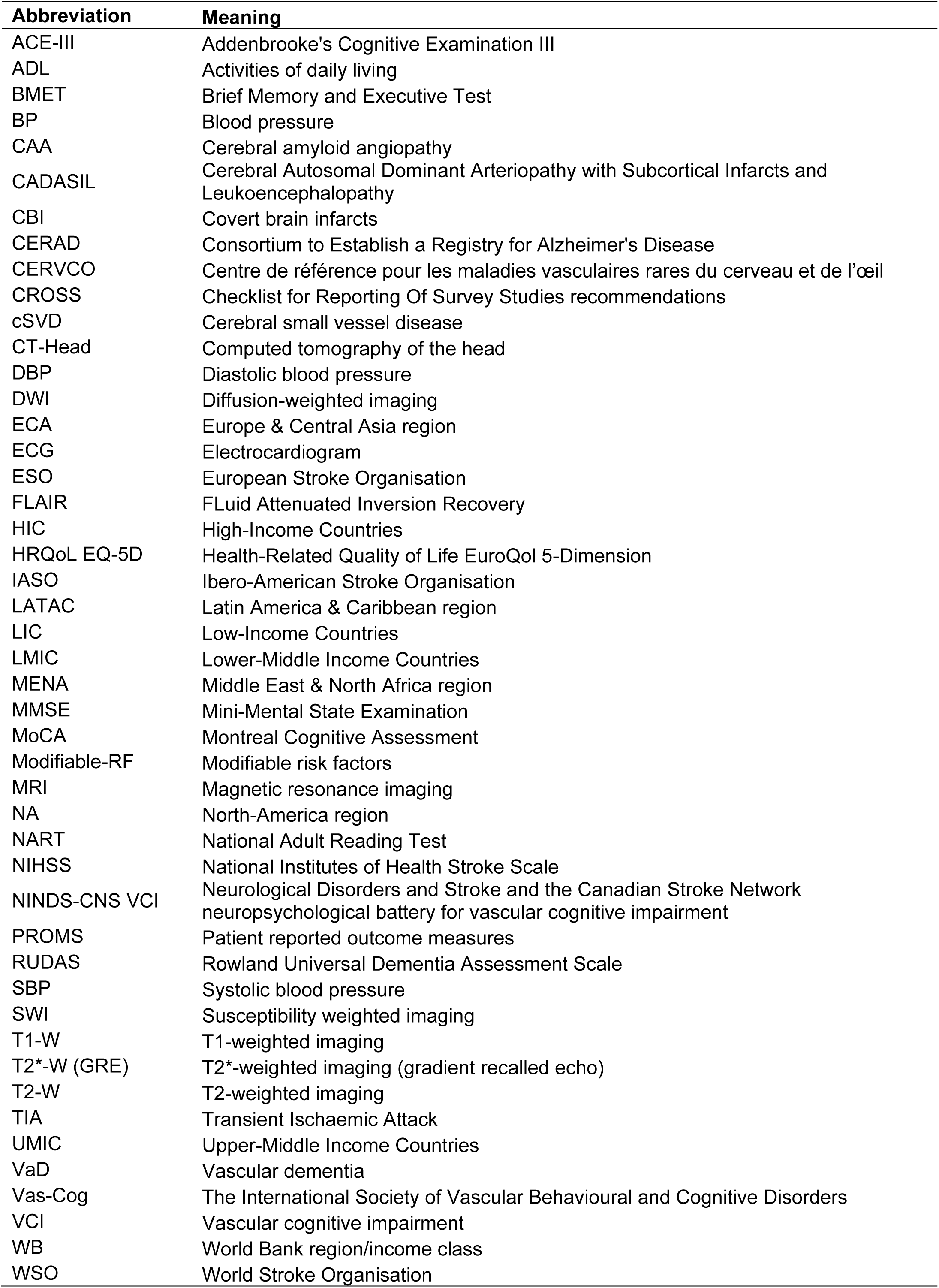

## Acknowledgements

We thank all the respondents of the international physician survey, and the Lothian NHS Cerebrovascular clinicians for their feedback in the pilot phase.

## Authorship

C.A.R, F.N.D., J.M.W.–Conceptualisation, design, results interpretation, writing, review, editing, manuscript review, approved final version of manuscript. C.A.R.–Acquisition of data, data analysis, writing original draft.

